# Diagnostic Utility of Endotracheal Aspirate Galactomannan for Invasive Pulmonary Aspergillosis in ICU Patients

**DOI:** 10.64898/2026.06.29.26356826

**Authors:** Ravi Kumar, Ankesh Gupta, Arvind Kumar, Sandeep Rao Kordcal, Upendra Baitha, Gagandeep Singh, Immaculata Xess, Karan Madan, Manish Soneja, Naveet Wig

## Abstract

**Background:** Invasive pulmonary aspergillosis (IPA) is a serious infection in critically ill patients. Galactomannan detection in endotracheal aspirates (ETA) has emerged as a promising non-invasive diagnostic method. This study evaluates the supportive diagnostic value of ETA galactomannan in ICU patients suspected to have IPA.

**Methods:** We conducted a prospective observational cohort study over two years, enrolling 120 patients in the medicine ICU at a tertiary care centre in India (January 2022 to October 2023). Patients aged over 14 years on mechanical ventilation for >48 hours meeting the entry criteria of the BM-AspICU algorithm were included. ETA galactomannan was measured and correlated with IPA classification.

**Results:** Of 120 patients, 37% (n=44) had probable IPA and 63% (n=76) were classified as colonisers or possible IPA. The optimal ETA galactomannan cut-off was 1.097, yielding sensitivity 72.73% (95% CI 57.2–85.0%), specificity 84.2% (95% CI 74.4–90.7%), PLR 4.86, NLR 0.35, and AUC 0.844

**Conclusion:** ETA galactomannan supports IPA diagnosis with favourable sensitivity and specificity. However, given the limitations of clinical scoring-based reference standards and the potential plateau in colonizer reduction at higher cut-offs, it should be integrated into a comprehensive diagnostic approach incorporating clinical, radiological, and microbiological criteria.

## Introduction

Aspergillosis refers to conditions caused by various *Aspergillus* species, such as *A. fumigatus, A. flavus, A. niger*, and *A. terreus*. Infection occurs through inhalation of airborne conidia, with invasive pulmonary aspergillosis (IPA) primarily affecting immunocompromised individuals, including those with hematologic malignancies, solid organ transplants, and critically ill patients [1-4]. Timely diagnosis is critical, as delayed treatment significantly increases mortality [5-7].

The incidence of invasive aspergillosis (IA) has grown globally due to increasing transplant recipients, broader use of immunosuppressive drugs, and improved ICU surveillance [8]. In India, invasive mycoses affect approximately 250,000 people annually [9]. Advances in diagnosis include detection of the *Aspergillus*-specific antigen galactomannan (GM), particularly in bronchoalveolar lavage (BAL) fluid and serum, and represents a major advance over conventional culture-based methods [10]. In ventilated patients, sampling from the lower respiratory tract has traditionally relied on bronchoscopic BAL; however, endotracheal aspirates (ETA) obtained by instillation of normal saline through the endotracheal tube with immediate aspiration have emerged as an accessible, less invasive alternative with direct access to the site of infection [11,12]. Studies have demonstrated that ETA galactomannan testing can guide further diagnostic workup and reduce the need for bronchoscopy [13,14]. This study evaluates ETA galactomannan as a supportive diagnostic tool for IPA in mechanically ventilated ICU patients.

## Methods

Between January 2022 and October 2023, patients admitted to the medicine ICU at a tertiary care centre were prospectively enrolled in this observational cohort study. The study was approved by the Institutional Ethics Committee of AIIMS, New Delhi (Ref. No. IECPG-326/27.04.22), and informed consent was obtained from legal representatives of all participants. Eligible patients were aged >14 years, had been on mechanical ventilation for >48 hours, and met the entry criteria of the BM-AspICU algorithm, a validated diagnostic framework for ICU patients that integrates host risk factors, mycological criteria, and clinical/radiological signs to classify patients as probable IPA or no IPA (coloniser/possible IPA) [15].

Clinical, radiological, and mycological data were recorded at recruitment. ETA was obtained by instillation of 20 mL sterile normal saline through the endotracheal tube with immediate aspiration within the first 24 hours. ETA samples were processed for galactomannan (Platelia™ *Aspergillus* EIA; Bio-Rad Laboratories) while bronchoalveolar lavage fluid was obtained for KOH stain, fungal culture/sensitivity, and *Aspergillus* PCR as well as galactomannan assay. Serum galactomannan and serum *Aspergillus* PCR were also obtained. For the galactomannan assay, an optical density index (ODI) ≥0.5 was used as the manufacturer-recommended positivity threshold. The sample size of 120 patients was determined by the number of eligible admissions over the study period (a formal power calculation was not performed). Patients were classified as probable IPA or no IPA (coloniser/possible IPA) per BM-AspICU criteria. (Figure 1)

**Figure 1.**
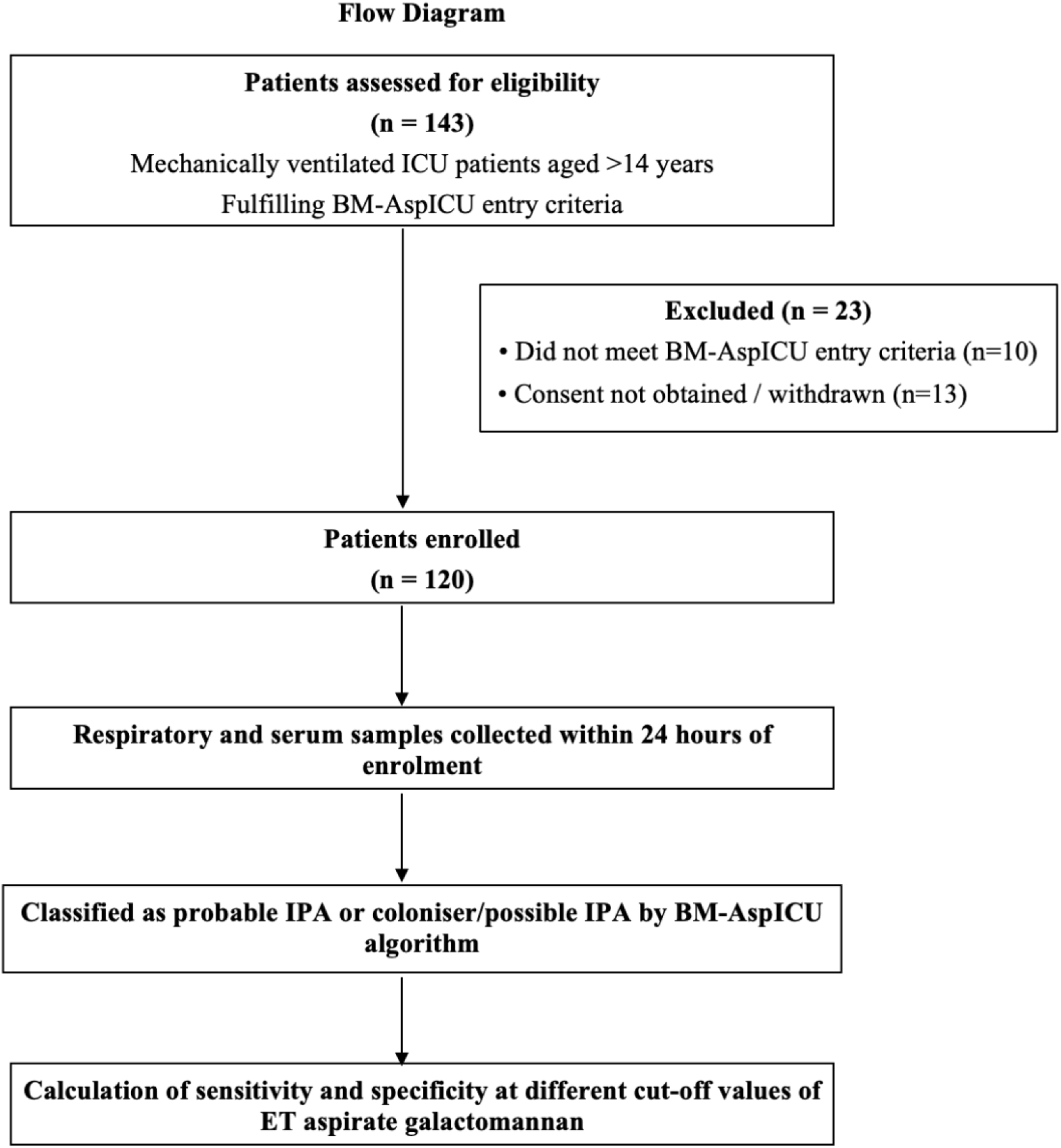
Study workflow. Critically ill ICU patients were screened against BM-AspICU entry criteria. Those meeting all inclusion/exclusion criteria with consent obtained were enrolled. Demographic, clinical, radiological, and mycological data were collected, and ETA galactomannan (plus serum galactomannan, BALF galactomannan, BAL and serum *Aspergillus* PCR, fungal culture) was obtained within 24 hours. Patients were classified as Probable IPA or coloniser/possible IPA per BM-AspICU algorithm, and diagnostic performance (sensitivity, specificity, PLR, NLR) at multiple cut-off values was calculated to determine the optimal ETA galactomannan threshold. (ICU= intensive care unit, BALF- Bronchoalveolar lavage fluid, IPA- invasive pulmonary aspergillosis, PCR- polymerase chain reaction).

Data were analysed using STATA version 14.0. Categorical variables were expressed as counts and percentages; quantitative variables as mean ± SD or median (IQR). Chi-square or Fisher’s exact test was used for categorical comparisons; Student’s t-test or Wilcoxon signed-rank test for continuous variables. A p-value <0.05 was considered statistically significant. Receiver operating characteristic (ROC) curve analysis was performed to evaluate the diagnostic performance of ETA galactomannan across a range of cut-off values (>0.5, >0.8, >1.0, >1.5, >2.0, >3.0). The optimal cut-off was identified using the Youden index (maximising sensitivity + specificity – 1). Sensitivity, specificity, positive and negative likelihood ratios (PLR, NLR), positive and negative predictive values (PPV, NPV), and area under the ROC curve (AUC) were calculated with 95% confidence intervals (CIs).

## Results

Of 143 patients screened, 120 met inclusion criteria. Forty-four (37%) were classified as probable IPA and 76 (63%) as colonisers or possible IPA. Mean age was 43.88 years (IPA group: 47.47 ± 16.1 vs. non-IPA: 41.80 ± 15.2 years; p=0.056). Gender distribution was similar between groups (55% male overall; p=0.939). Median hospital stay was comparable (probable IPA: 20.5 days, IQR 9.5–34 vs. non-IPA: 22.5 days, IQR 12–32.5; p=0.74). Comorbidities were distributed similarly across groups. (Table 1). Mortality was significantly higher in the probable IPA group (86.36% vs. 53.95%; p=0.001).

**Table 1:** Baseline, clinical, and radiological characteristics of the study population.

| **Parameter (n=120)** | **Probable IPA (n=44)** | **Coloniser/Possible IPA (n=76)** | **p-value** |
| --- | --- | --- | --- |
| Age (years), mean ± SD | 47.5 ± 16.1 | 41.8 ± 15.2 | 0.058 |
| Male sex, n (%) | 24 (54.6%) | 42 (55.2%) | 0.939 |
| Hospital stay (days), median (IQR) | 20.5 (9.5, 34) | 22.5 (12, 32.5) | 0.74 |
| Comorbidities, n (%) |  |  |  |
| Diabetes mellitus | 13 (29.55%) | 18 (23.68%) | 0.480 |
| Obstructive airway disease | 4 (9.09%) | 4 (5.26%) | 0.417 |
| Chronic liver disease | 6 (13.64%) | 12 (15.78%) | 0.750 |
| Chronic kidney disease | 17 (38.63%) | 21 (27.63%) | 0.212 |
| Mortality, n (%) | 38 (86.36%) | 41 (53.95%) | 0.001 |
| Clinical features |  |  |  |
| Fever | 38 (86.3%) | 53 (69.7%) | 0.04 |
| Haemoptysis | 0 | 0 | n/a |
| Worsening on mechanical ventilation | 15 (34.09%) | 18 (23.68%) | 0.219 |
| Radiological features |  |  |  |
| Air crescent sign | 0 | 0 | n/a |
| Cavity | 4 (9.09%) | 4 (5.26%) | 0.668 |
| CT nodules | 22 (50%) | 30 (39.47%) | 0.070 |
| X-ray/CT consolidation | 31 (70.45%) | 42 (55.26%) | 0.100 |
| Pleural effusion | 18 (40.9%) | 26 (34.21%) | 0.463 |
| Tree-in-bud/wedge infiltrate | 4 (9.09%) | 6 (7.89%) | 0.463 |

IPA: invasive pulmonary aspergillosis; IQR: interquartile range; CT: computed tomography; n/a: not applicable.

Fever was significantly more common in probable IPA patients (86.3% vs. 69.7%, p=0.04), while other clinical and radiological findings, including worsening respiratory status, cavitation, consolidation, pleural effusion, and CT nodules were numerically more frequent but did not differ significantly between groups.

Microbiologically, *Aspergillus* species were isolated in 13.3% (n=16) of patients: *A. fumigatus* (n=9), *A. flavus* (n=6), and *A. terreus* (n=1).

The diagnostic performance of ETA galactomannan across a range of cut-off values is detailed in Table 2. Sensitivity decreased progressively with rising cut-offs, from 93.2% (95% CI 81.3–98.6%) at >0.5 to 29.5% (95% CI 16.8–45.2%) at >3.0, whilst specificity improved from 46.1% (95% CI 34.5–57.9%) at >0.5 to 96.1% (95% CI 88.9–99.2%) at >3.0. Using the Youden index, the optimal cut-off was identified as 1.097, yielding sensitivity 72.73% (95% CI 57.2–85.0%), specificity 84.2% (95% CI 74.4–90.7%), PLR 4.86, NLR 0.35, PPV 68.1% (95% CI 52.9–80.9%), NPV 83.6% (95% CI 73.0–91.2%), and AUC 0.844 (Figure 2), indicating favourable overall diagnostic performance.

**Table 2:** Diagnostic performance of endotracheal aspirate (ETA) galactomannan at different cut-off values (95% CI).

| Parameter | >0.5 | >0.8 | >1.0 | >1.5 | >2.0 | >3.0 |
| --- | --- | --- | --- | --- | --- | --- |
| Sensitivity | 93.2% (81.3–98.6%) | 84.1% (72.8–91.8%) | 72.73% (57.2–85.0%) | 52.3% (36.7–67.5%) | 38.6% (24.4–54.5%) | 29.5% (16.8–45.2%) |
| Specificity | 46.1% (34.5–57.9%) | 69.74% (55.8–80.8%) | 80.3% (69.5–88.5%) | 90.8% (81.9–96.2%) | 94.7% (87.1–98.5%) | 96.1% (88.9–99.2%) |
| PLR | 1.73 (1.38–2.16) | 2.77 (1.85–4.17) | 3.68 (2.26–6.00) | 5.68 (2.65–12.14) | 7.34 (2.64–20.40) | 7.48 (2.26–24.83) |
| NLR | 0.15 (0.05–0.45) | 0.22 (0.11–0.45) | 0.34 (0.21–0.56) | 0.53 (0.38–0.72) | 0.65 (0.51–0.82) | 0.73 (0.60–0.89) |
| PPV | 50% (38.7–61.3%) | 62.7% (49.1–75%) | 68.1% (52.9–80.9%) | 76.7% (57.7–90.1%) | 81.0% (58.1–94.6%) | 81.3% (54.4–96.0%) |
| NPV | 92.1% (78.6–98.3%) | 88.5% (77.8–95.3%) | 83.6% (73–91.2%) | 76.7% (66.6–84.9%) | 72.7% (62.9–81.2%) | 70.2% (60.4–78.8%) |

PLR: positive likelihood ratio; NLR: negative likelihood ratio; PPV: positive predictive value; NPV: negative predictive value.

**Figure 2.**
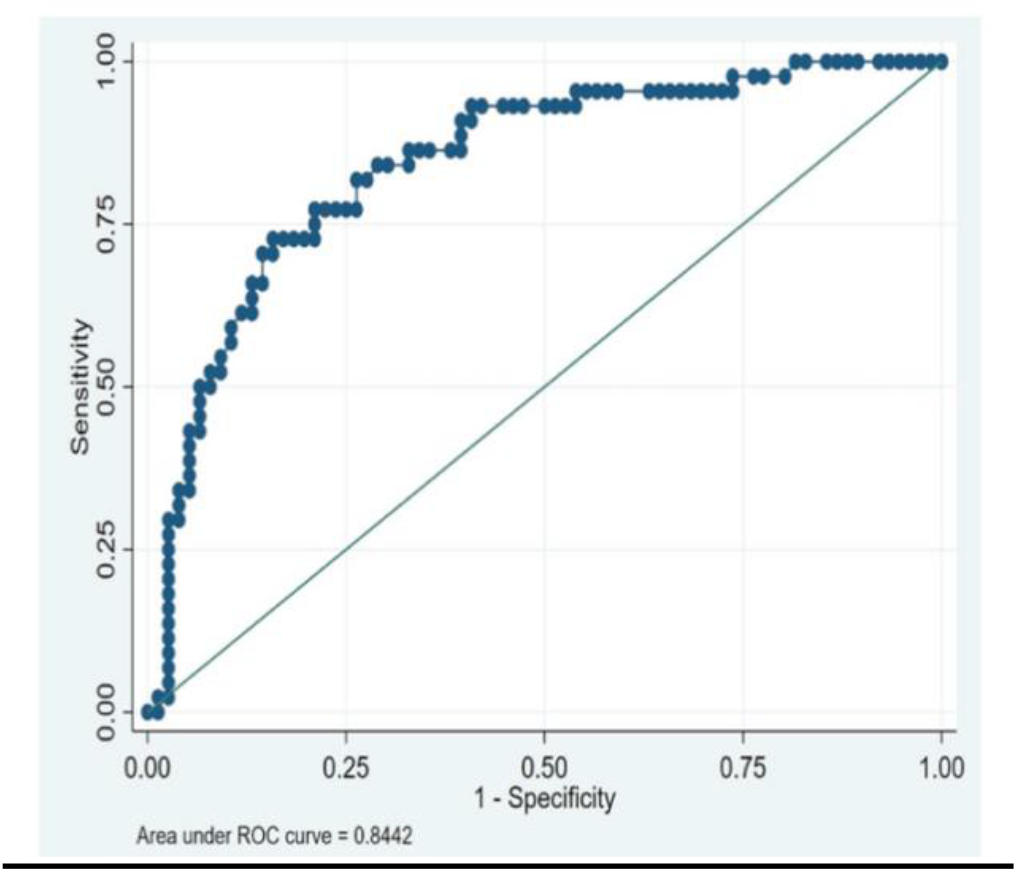
ROC curve of endotracheal aspirate galactomannan, showing the optimal cut-off value of 1.097 with sensitivity 72.73% (95% CI 57.2–85.0%), specificity 84.2% (95% CI 74.4–90.7%), PLR 4.86, NLR 0.35, and AUC 0.844. (ROC: Receiver Operating Characteristic curve)

The relationship between ETA galactomannan cut-off values and the proportion of colonisers was also examined (Table 3, Figure 3). At a cut-off of >0.5, 50% of cases were classified as colonisers. This proportion fell to 38.33% at >0.8 and to 31.91% at >1.0. Further increases in the cut-off yielded diminishing reductions: 23.3% at >1.5, 19.0% at >2.0, and 18.75% at >3.0. This pattern indicates a ceiling effect/plateau beyond a cut-off of >1.5, beyond which further increases in threshold provide little additional discrimination between true infection and colonisation.

**Table 3:** Proportion of colonisers at different ETA galactomannan cut-off thresholds.

| ETA Galactomannan Cut-off | Proportion of Colonisers (%) |
| --- | --- |
| >0.5 | 41/82 (50.0%) |
| >0.8 | 23/60 (38.33%) |
| >1.0 | 15/47 (31.91%) |
| >1.5 | 7/30 (23.3%) |
| >2.0 | 4/21 (19.0%) |
| >3.0 | 3/16 (18.75%) |

**Figure 3.**
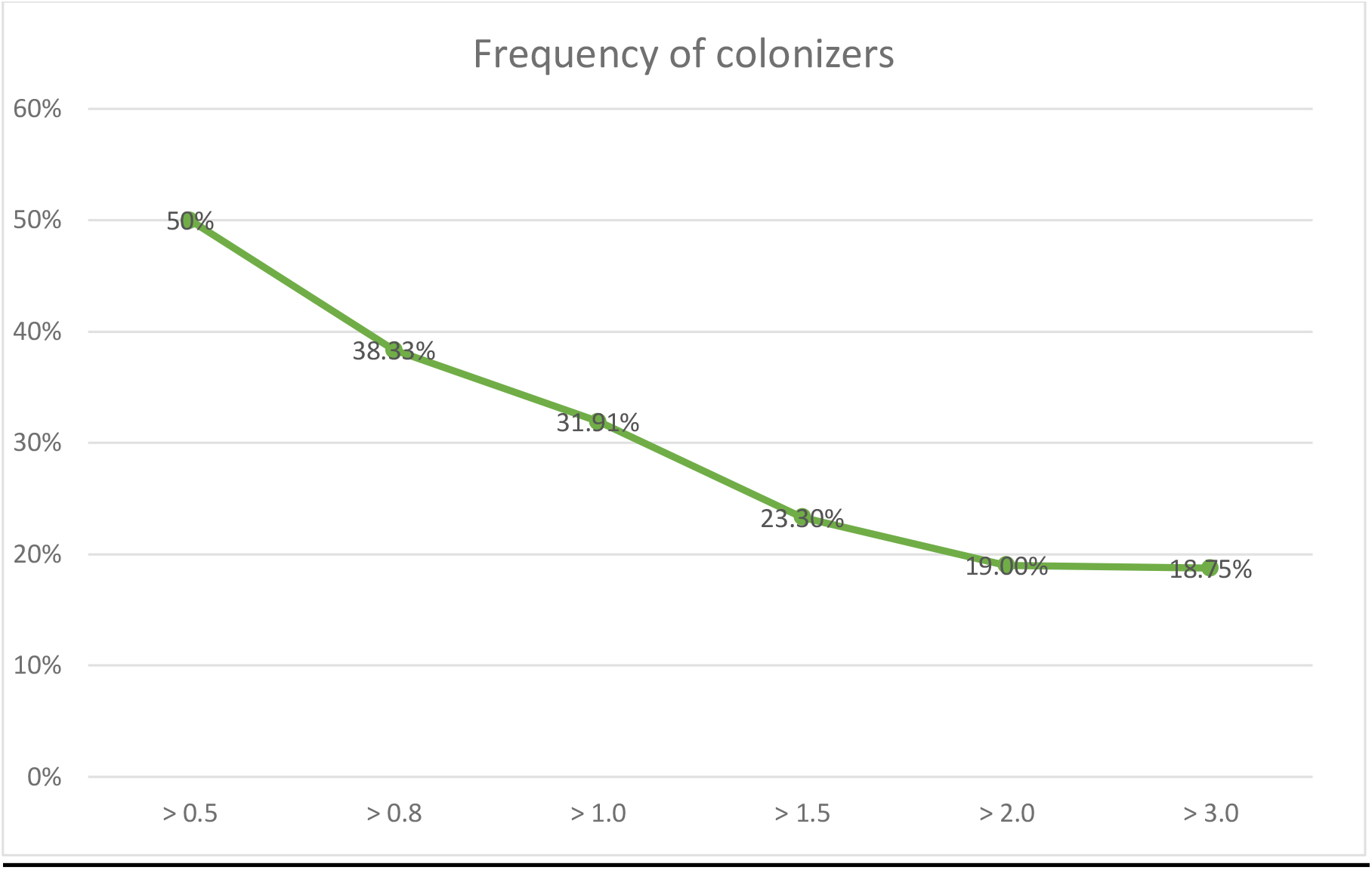
Proportion of colonizers at different endotracheal aspirate galactomannan cut-off thresholds, illustrating reduction in colonizer proportion at cut-offs >1.5.

## Discussion

This prospective study evaluated ETA galactomannan as a supportive diagnostic tool for IPA in 120 mechanically ventilated ICU patients. At the optimal cut-off of 1.097, sensitivity was 72.73% (95% CI 57.2–85.0%) and specificity was 84.2% (95% CI 74.4– 90.7%) with an AUC of 0.844, indicating favorable diagnostic performance within a comprehensive clinical framework. These figures compare favourably with published data: Roman-Montes et al. [13] reported ETA galactomannan sensitivity of 87% and specificity of 76% in the specific context of COVID-19-associated pulmonary aspergillosis (CAPA), a distinct but related entity, while Dichtl et al. [14] demonstrated that ETA galactomannan testing could effectively guide subsequent bronchoscopic sampling decisions. The identified cut-off of 1.097 is also consistent with the galactomannan index of ≥1.0 recommended for BAL in the ESCMID/ECMM/ERS guidelines [16], supporting ETA as a viable, less invasive non bronchoscopic adjunct in patients where bronchoscopy carries significant procedural risk.

Comorbidities, including chronic kidney disease, diabetes mellitus, and obstructive airway disease, were distributed similarly across groups and did not differ significantly. The markedly elevated mortality in the probable IPA group (86.36% vs. 53.95%; p=0.001) underscores the severity of IPA in the ventilated ICU patient and the critical importance of early biomarker-guided diagnosis.

Among clinical and radiological features, fever was the only finding reaching statistical significance (86.3% vs. 69.7%; p=0.04), though concomitant VAP and multidrug-resistant Gram-negative bacteraemia limited its specificity for IPA. CT nodules showed a trend toward significance (p=0.070); Microbiological yield was low (13.3% culture positivity), consistent with known difficulties in *Aspergillus* culture from respiratory specimens.

The behaviour of ETA galactomannan across a range of cut-off thresholds warrants detailed consideration. At the manufacturer-recommended cut-off of >0.5, sensitivity was high (93.2%) but specificity was poor (46.1%), making it most useful as a rule-out test: a negative result at this threshold substantially reduces the probability of IPA (NLR 0.15). As the cut-off was raised, specificity improved progressively, reaching 96.1% at >3.0, at the cost of sensitivity, which fell to 29.5%. The Youden-optimal cut-off of 1.097 balanced these trade-offs, yielding sensitivity 72.73%, specificity 84.2%, and an AUC of 0.844, with a clinically useful PLR of 4.86, indicating that a positive result at this threshold meaningfully raises post-test probability for IPA. A plateau was evident beyond cut-offs of >1.5, where further increases in threshold produced diminishing reductions in coloniser misclassification (from 23.3% at >1.5 to 18.75% at >3.0). This plateau underscores that galactomannan thresholding alone cannot reliably distinguish true IPA from airway colonisation, and that the optimal cut-off should be selected according to clinical context: a lower threshold (>0.5) favours sensitivity and is appropriate for screening high-risk patients or guiding further workup, while a higher threshold (>1.5 or above) may be preferable when greater specificity is required to justify antifungal initiation.

These findings reinforce that ETA galactomannan functions best as a first-line adjunct within a stepwise diagnostic strategy rather than as a standalone test. When ETA galactomannan is positive at or above the optimal threshold, further diagnostic workup should be systematically pursued. Fungal cultures, though limited by low sensitivity (13.3% in this cohort), retain value for species identification and antifungal susceptibility profiling, particularly given the emergence of azole-resistant *A. fumigatus* globally. Advanced molecular diagnostics including metagenomic next-generation sequencing (mNGS) are increasingly complementing conventional methods in resource-adequate settings, enabling pathogen-agnostic detection and circumventing the sensitivity limitations of culture-based approaches [17].

The principal strength of this study lies in its prospective design, which reduces the risk of selection and information bias inherent in retrospective analyses. All patients underwent a comprehensive mycological workup enabling a robust multi-modal diagnostic assessment. The study is also among the few from a resource-limited South Asian ICU setting, where invasive procedures such as BAL are not always feasible, underscoring the practical value of ETA galactomannan as a frontline diagnostic adjunct.

### Limitations

Several limitations should be considered when interpreting these findings. First, the absence of histological confirmation means that classification relied on BM-AspICU clinical criteria rather than a microbiological gold standard; proven IPA cases were not available in this cohort, which constrains the formal diagnostic validation. Second, no formal power calculation was performed; the sample size was dictated by the number of eligible admissions, which may have underpowered subgroup analyses. Third, a small subset of patients underwent non-bronchoscopic mini-bronchoalveolar lavage (mini-BAL) for *Aspergillus* PCR and fungal culture rather than standard bronchoscopic BAL, owing to the limited availability of bronchoscopy in a resource-constrained ICU setting and comparable performance [12]. Fourth, the single-centre design at a tertiary referral hospital limits generalisability to other ICU settings. Finally, secondary bacterial co-infections such as VAP and multidrug-resistant Gram-negative bacteraemia may have confounded radiological and clinical findings attributed to IPA.

## Conclusion

ETA galactomannan demonstrates favourable sensitivity and specificity as a supportive tool for diagnosing IPA in mechanically ventilated ICU patients. Given the methodological constraints of clinical criteria-based classification and the plateau in colonizer reduction at higher cut-offs, this biomarker should be integrated into a comprehensive diagnostic strategy encompassing clinical evaluation, radiological findings, and microbiological evidence. Further studies validating ETA galactomannan against histologically confirmed cases are warranted.

## Data Availability

All data produced in the present study are available upon reasonable request to the authors

## Acknowledgements

Department of Medicine, AIIMS, New Delhi; Department of Microbiology, AIIMS, New Delhi; Department of Pulmonary Medicine, AIIMS, New Delhi.

## Author Contributions

Ravi Kumar and Ankesh Gupta: data collection, laboratory analysis, and manuscript drafting. Arvind Kumar: study conception, design, supervision, and critical revision of the manuscript. Sandeep Rao Kordcal : manuscript drafting and revision. Gagandeep Singh and Immaculata Xess: mycological analyses and interpretation. Upendra Baitha, Manish Soneja, Karan Madan and Naveet Wig: study oversight and critical revision. All authors approved the final version for submission.

## Data Availability Statement

The data that support the findings of this study are available from the corresponding author upon reasonable request, subject to institutional data governance and patient confidentiality requirements.

## References

[1] Fosses Vuong M, Hollingshead CM, Waymack JR. Aspergillosis. In: StatPearls [Internet]. Treasure Island (FL): StatPearls Publishing; 2023.

[2] Ledoux MP, Guffroy B, Nivoix Y, et al. Invasive pulmonary aspergillosis. Semin Respir Crit Care Med. 2020; 41(1): 80–98.

[3] Tejerina EE, Abril E, Padilla R, et al. Invasive aspergillosis in critically ill patients: an autopsy study. Mycoses. 2019; 62(8): 673–679.

[4] Pardo E, Lemiale V, Mokart D, et al. Invasive pulmonary aspergillosis in critically ill patients with hematological malignancies. Intensive Care Med. 2019; 45(12): 1732–1741.

[5] McNeil MM, Nash SL, Hajjeh RA, et al. Trends in mortality due to invasive mycotic diseases in the United States, 1980-1997. Clin Infect Dis. 2001; 33(5): 641–647.

[6] Henao-Martínez AF, Corbisiero MF, Salter I, et al. Invasive pulmonary aspergillosis real-world outcomes: clinical features and risk factors associated with increased mortality. Med Mycol. 2023; 61(8): myad074.

[7] Shi C, Shan Q, Xia J, et al. Incidence, risk factors and mortality of invasive pulmonary aspergillosis in patients with influenza: a systematic review and meta-analysis. Mycoses. 2022; 65(2): 152–163.

[8] Vallabhaneni S, Benedict K, Derado G, et al. Trends in hospitalizations related to invasive aspergillosis and mucormycosis in the United States, 2000–2013. Open Forum Infect Dis. 2017; 4(1): ofw268.

[9] Ray A, Aayilliath KA, Banerjee S, et al. Burden of serious fungal infections in India. Open Forum Infect Dis. 2022; 9(12): ofac603.

[10] Cuenca-Estrella M, Bassetti M, Lass-Florl C, et al. Detection and investigation of invasive mould disease. J Antimicrob Chemother. 2011; 66(Suppl 1): i15–i24.

[11] Khilnani GC, Arafath TK, Hadda V, et al. Comparison of bronchoscopic and nonbronchoscopic techniques for diagnosis of ventilator-associated pneumonia. Indian J Crit Care Med. 2011; 15: 16–23.

[12] Agarwal A, Malviya D, Harjai M, et al. Comparative evaluation of the role of nonbronchoscopic and bronchoscopic techniques of distal airway sampling for the diagnosis of ventilator-associated pneumonia. Anesth Essays Res. 2020; 14: 434–440.

[13] Román-Montes CM, Bojorges-Aguilar S, Díaz-Lomelí P, et al. Tracheal aspirate galactomannan testing in COVID-19-associated pulmonary aspergillosis. Front Fungal Biol. 2022; 3: 855914.

[14] Dichtl K, Barry R, Angstwurm MWA, Suerbaum S, Wagener J. Performance of galactomannan testing from endotracheal aspirate to guide bronchoalveolar lavage in the diagnosis of invasive aspergillosis. Infection. 2023;51(3):769–774. doi:10.1007/s15010-023-01985-1

[15] Hamam J, Navellou JC, Bellanger AP, et al. New clinical algorithm including fungal biomarkers to better diagnose probable invasive pulmonary aspergillosis in ICU. Ann Intensive Care. 2021;11(1):41. Published 2021 Mar 8. doi:10.1186/s13613-021-00827-3

[16] Ullmann AJ, Aguado JM, Arikan-Akdagli S, et al. Diagnosis and management of Aspergillus diseases: executive summary of the 2017 ESCMID-ECMM-ERS guideline. Clin Microbiol Infect. 2018; 24(Suppl 1): e1–e38.

[17] Plazola CE, Rehman A, Morel M, et al. Aspergillus fumigatus endocarditis in an immunocompetent host aided by metagenomic next-generation sequencing assay: case report and literature review. Infez Med. 2025; 33(2): 226–232.

